# A Cross-Dataset Audit of Discrimination and Conformal Prediction Coverage in Keystroke-Based Parkinson’s Disease Classification

**DOI:** 10.64898/2026.11.04.26366890

**Authors:** Haotian Guang, Zijuan Zhang, Jiale Qu, Jiaqiang Wu, Jiayuan Ma

**Affiliations:** School of Life Sciences, Inner Mongolia University, Hohhot, China

**Keywords:** Parkinson’s disease, Keystroke dynamics, Conformal prediction, Digital biomarkers, Distribution shift, Machine learning

## Abstract

Typing dynamics are being investigated as a digital behavioral marker for Parkinson’s disease (PD), but the cross-dataset reliability of typing-based classifiers remains uncertain. We evaluated four public datasets (475 participants; 270 with PD and 205 controls) across 12 transfer directions using split conformal prediction targeting 90% marginal coverage. In all three primary directions, coverage was below the 0.90 target and the one-sided tests remained significant after Holm correction: 0.598 (95% CI, 0.483–0.704) for Tappy→MIT, 0.850 (0.788–0.899) for MIT→Tappy, and 0.732 (0.684–0.777) for clinical→self-reported transfer. Discrimination and coverage did not consistently change together: LightGBM had lower AUC than logistic regression in all 12 directions but higher coverage in 11, with larger prediction sets. Class-conditional coverage varied by target cohort: PD-conditional coverage was 0.214 for Tappy→MIT, whereas healthy-control coverage was 0.700 for MIT→Tappy and 0.627 for clinical→self-reported transfer. Recalibration with 9 and 18 labeled target participants raised the lower confidence bound above 0.80 in the latter two directions; in Tappy→MIT, it remained below 0.80 even with 25 labels. Weighted conformal prediction yielded coverage of 0.918–1.000 without target labels but returned the full label set for 86%–100% of participants. Stress tests did not reproduce the observed coverage patterns, and black-box shift estimation did not support pure label shift as the dominant explanation for Tappy→MIT. Evaluating typing dynamics as a digital marker for PD requires cross-dataset assessment of discrimination, marginal and class-conditional coverage, and recovery costs. Clinical screening utility has not been established.

## 1. Introduction

In recent years, machine-learning classifiers based on typing dynamics (keystroke dynamics) have been used for Parkinson’s disease (PD) screening and have demonstrated promising discriminative performance |1,5,13,15]. Publicly available datasets have facilitated the continued development and validation of related models over the past decade. The performance of existing machine-learning classifiers based on typing dynamics for PD has been evaluated primarily in terms of discrimination, typically measured by the area under the receiver operating characteristic curve (AUC) [2,12,23]. However, when a model is transferred from its development dataset to a new dataset, distribution shift may arise from changes in input feature distributions, participant composition, data acquisition devices, task designs, and sources of diagnostic labels. Therefore, the evaluation of cross-dataset deployment needs to consider both model discrimination and the reliability of predictive uncertainty.

Conformal prediction (CP) provides a model-agnostic framework with growing clinical adoption [7,9,26] for quantifying predictive uncertainty [4]. It characterizes the reliability of uncertainty quantification by generating prediction sets and evaluating their coverage. When the calibration data and the target data satisfy exchangeability, split conformal prediction provides a finite-sample guarantee for marginal coverage, with the true label being included in the prediction set with probability at least 1-*α*. However, cross-dataset deployment may violate exchangeability [6], rendering the standard coverage guarantee inapplicable to the target data and potentially causing the empirical coverage to deviate from its nominal level. Changes in conformal coverage across publicly available PD typing datasets, and the relationship between coverage degradation and degradation in discrimination, have not yet been systematically investigated [20,25] through cross-dataset audits.

AUC measures a model’s ability to discriminate between classes, whereas conformal coverage measures the frequency with which prediction sets contain the true label. The two metrics have distinct statistical interpretations, and their changes are not necessarily coupled under distribution shift [18,22]. This raises a direct empirical question: when PD typing classifiers are deployed across datasets, do coverage degradation and discrimination degradation vary in tandem across different transfer directions? We hypothesize that coverage degradation and discrimination degradation exhibit different patterns of change under distribution shift and should therefore be treated as distinct dimensions of deployment reliability. If this hypothesis holds, external evaluation based solely on AUC cannot fully characterize changes in predictive reliability.

In this study, we conducted an audit of conformal coverage for PD typing classifiers deployed across datasets, covering all 12 ordered transfer directions among four publicly available datasets. We compared AUC and marginal coverage across transfer directions to examine whether coverage degradation and discrimination degradation occurred in tandem and characterized class-specific coverage failure patterns through class-conditional coverage diagnostics. We then investigated the potential sources of coverage degradation using controlled stress tests and evaluated the amount of target-domain labeling required to restore coverage, as well as the associated costs in prediction set-efficiency, under different calibration strategies.

## 2. Methods

### 2.1 Data sources

The data used in this study were obtained from four publicly available datasets: MIT-CSXPD (the neuroQWERTY project; PhysioNet, nqmitcsxpd v1.0.0 [13]), Tappy (PhysioNet, tappy v1.0.0 [1]), TyPD (i-PROGNOSIS touchscreen typing data; Zenodo record 2571623 [14]), and Online English (OE; OSF repository ew34b [11]). These datasets are heterogeneous in terms of diagnostic information sources, typing platforms, devices, and task designs. MIT-CSXPD and TyPD provide clinical diagnostic labels, whereas Tappy and OE use self-reported diagnostic labels. The data collection set-tings span clinical environments, personal devices, and web-based platforms. The source publications report an accrual period only for Tappy (July 2016 to March 2017) [1]; MIT-CSXPD, TyPD, and OE do not report accrual dates.

### 2.2. Cohort construction and data cleaning

Digraphs (ordered pairs of consecutive keystrokes) were used as the basic units for feature construction. Data cleaning and filtering were performed without reference to participants’ diagnostic labels, relying solely on the quality of the typing data. Digraphs were first extracted from the raw keystroke records, retaining valid events that passed parsing and validation checks and had complete key-down and key-up timestamps. On this basis, uniform thresholds for key-hold duration and inter-keystroke interval were applied across all datasets to remove anomalous data, followed by additional task-specific rules for the three datasets that used fixed-text transcription tasks. Next, a continuous period of typing was defined as a session, and session segmentation criteria were established for each dataset. For the MIT clinical dataset, only sessions from each participant’s first visit were retained. Finally, all valid digraphs were aggregated for each participant, and participants with fewer than 300 digraphs were excluded. Detailed filtering rules, the number of samples excluded at each step, and the final cohort composition are provided in Supplementary Tables S1 and S4.

### 2.3. Feature construction and prediction models

All features were aggregated at the participant level, with all valid digraphs from each participant used for feature calculation. Features were constructed at three levels of complexity: L1 calculated distributional statistics (mean, median, standard deviation, 10th percentile, and 90th percentile) for each of the three temporal measures, hold (key press duration), latency (time from the release of the previous keystroke to the press of the current keystroke), and flight (time interval between two consecutive keystrokes), yielding 15 features available across all datasets; L2 added mean features for left- and right-hand conditions to L1 and was supported only by MIT, Tappy, and OE; L3 aggregated mean latency and flight values according to hand transition types between adjacent digraphs and included the proportion of transitions between hands, with this level supported only by MIT and OE. For each transfer direction, the highest level supported by both the source and target datasets was used, and results stratified by feature level were reported as sensitivity analyses.

The primary model was *l*2-regularized logistic regression (feature standardization was performed only within the training set), the comparator model was LightGBM, a gradient boosting decision-tree framework, with a shallow-tree configuration (maximum depth of 3; minimum number of sam-ples per leaf set to one-quarter of the training set), and the parameters were not adjusted thereafter. Both models were evaluated within the MIT source domain using internal splits with 50 random seeds, yielding mean test AUCs of 0.787 and 0.726, respectively. This evaluation did not involve target data; its results also served as the development-domain reference for ΔAUC.

### 2.4. Conformal prediction framework

Split conformal prediction (Split CP) was used as the coverage audit framework, with the Least Ambiguous Set-Valued Classifier (LAC) score [4] as the nonconformity score, defined as one minus the model’s predicted probability for the true label. The lower the predicted probability for the true label, the higher the LAC score, indicating greater nonconformity between the true label and the prediction model.

The calibration set was sampled from the source cohort using class-stratified sampling and accounted for 30% of the source cohort. The conformal threshold was calculated jointly using all scores in the calibration set, targeting a marginal coverage of 1−α. In this study, α = 0.10, corresponding to a nominal coverage of 90%. The conformal threshold was set to the k-th smallest LAC score in the calibration set, where k = ⌈(n_cal_+1)(1−α)⌉ and n_cal denotes the calibration set size. A candidate label was included in the prediction set if its LAC score did not exceed this threshold, and the prediction set consisted of all classes when k > n_cal_. The primary analysis used a single split corresponding to the first of the 50 random seeds, and the mean results across the 50 seeds were used only for descriptive analysis. Class-conditional coverage was used as a diagnostic metric for failure modes. Under the exchangeability assumption, the theoretical guarantee of Split CP is marginal coverage of 1−α.

We evaluated three coverage recovery strategies to assess the feasibility and cost of restoring coverage. For target-domain recalibration, 5%, 10%, 20%, or 30% of labeled target-domain samples were selected using class-stratified sampling to recalculate the threshold. Weighted conformal prediction (weighted CP) used density ratios between the target and source domains as calibration weights, estimated as the odds output of a logistic-regression domain classifier trained on the source training samples and all target samples with class-balancing weights under an equal-domain prior. Following the weighted conformal prediction framework for covariate shift, the weights were truncated at the pooled 99th percentile (p99), which forfeits the formal weighted marginal-coverage guarantee that holds without truncation. Protocol-conditional recalibration calculated calibration quantiles separately for each acquisition protocol, setting the target marginal coverage within each protocol group to 1−α, conditional on exchangeability holding within each group.

### 2.5. Statistical analysis

All inferential analyses used the participant as the unit of analysis: each participant’s session data and valid digraph events were aggregated beforehand, and each participant contributed one binary (0/1) observation to target-domain coverage. The primary audit metric in this study was marginal coverage for each transfer direction, and inferential tests were conducted only for three primary transfer directions: Tappy→MIT, MIT→Tappy, and the pooled transfer direction from the clinical datasets (MIT+TyPD) to the self-reported datasets (Tappy+OE). The results for all 12 ordered transfer directions among the four datasets formed a descriptive audit matrix. Marginal coverage in each primary direction was assessed using a one-sided binomial test, with the null hypothesis that the true coverage was ≥ 0.90 and a nominal significance level of α = 0.10. The Holm correction was applied to control the family-wise error rate at 0.10 across this family of tests. The binomial tests evaluate coverage conditional on the fixed training and calibration split; the 50-seed summaries are descriptive and do not support inference about split-averaged coverage, because the splits reuse the same participants.

Discrimination was measured using participant-level AUC, with the mean and standard deviation across 50 random seeds reported for each direction. The development-domain reference was the mean AUC from the source-domain evaluation described above (0.787 and 0.726), and ΔAUC was defined as the difference between each direction’s mean AUC and this reference. This MIT-based benchmark was shared across all directions; ΔAUC therefore measures deviation from a fixed development-domain reference rather than the source-specific change in discrimination caused by transfer. The central hypothesis of this study was that coverage degradation and discrimination degradation exhibit different patterns of change under distribution shift. It was assessed descriptively at the direction level by jointly interpreting ΔAUC and the deviation of marginal coverage from the nominal level of 0.90 as continuous quantities; patterns consistent with the hypothesis were those in which one dimension departed from its reference while the other did not.

The operational audit further compared unadjusted 95% Clopper − Pearson intervals with the 0.80 audit threshold, with the rules applied sequentially: directions with an upper confidence bound below 0.90 were classified as Incompatible; otherwise, directions with a lower confidence bound of at least 0.80 were classified as Compatible; all remaining directions were classified as Undetermined.

### 2.6. Recovery and stress-test analyses

The recovery-cost analysis evaluated target-domain recalibration (labeling budgets of 5/10/20/30%), weighted CP, and protocol-conditional recalibration, each against unmodified split CP, and reported the number of target-domain labels, coverage, PD-conditional coverage (coverage among participants with PD), mean set size, singleton rate, and the full-label-set output rate (the proportion of participants receiving all labels; rejection rate). Costs were measured along two dimensions: the amount of target-domain labeling as information cost, and mean set size, singleton rate, and full-label-set output rate as prediction-set efficiency costs. Recovery outcomes were assessed according to the audit thresholds and confidence interval-based decision rules described above.

The three stress tests evaluated whether candidate shift mechanisms could produce coverage or distributional characteristics consistent with the observed results. First, in a synthetic dose-response experiment, participant-level labels were flipped at doses of 10%, 20%, and 30% for the PD class, the HCclass, or both, and the results were compared with the theory of dispersive label noise 3; the theory’s requirement that the post-flipping calibration-score distribution first-order stochastically dominate the clean distribution was verified in all theory-aligned combinations. Second, three manipulation scenarios were implemented in the MIT→Tappy direction: resampling the source cohort to match the target prevalence, bidirectional label flipping at unchanged prevalence, and their combination, with practical equivalence assessed using tolerances of ±5 percentage points for coverage and ±0.3 for mean set size. Finally, black-box shift estimation (BBSE) [24], the confusion-matrix estimator of class prevalences under prior probability shift, was used to estimate the target prevalence from the class-conditional confusion matrix of the source-trained classifier, under the label shift assumption that P(x | y) remains unchanged.

## 3. Results

### 3.1. Analytical cohort

The final analytical cohort comprised 475 participants across the four datasets (270 with PD and 205 healthy controls (HC); MIT 82, Tappy 173, Ty PD 27, and OE 193; Supplementary Table S4).

### 3.2. Cross-dataset marginal coverage audit

The three primary transfer directions were evaluated on 82, 173, and 366 target participants, respectively (Supplementary Table S4). After Holm correction, the one-sided tests for all three directions rejected the hypothesis that the true coverage was ≥0.90 (Figure 1; Supplementary Table S5). Tappy→MIT 0.598 [0.483, 0.704] (adjusted p = 1.2×10-12), MIT→Tappy 0.850 [0.788, 0.899] (adjusted p=0.023), and clinical pool (MIT+Ty PD)→self-reported (Tappy+OE) 0.732 [0.684, 0.777] (adjusted p = 2.5×10-19). All three directions were classified as Incompatible (Supplementary Table S5).

**Figure 1.**
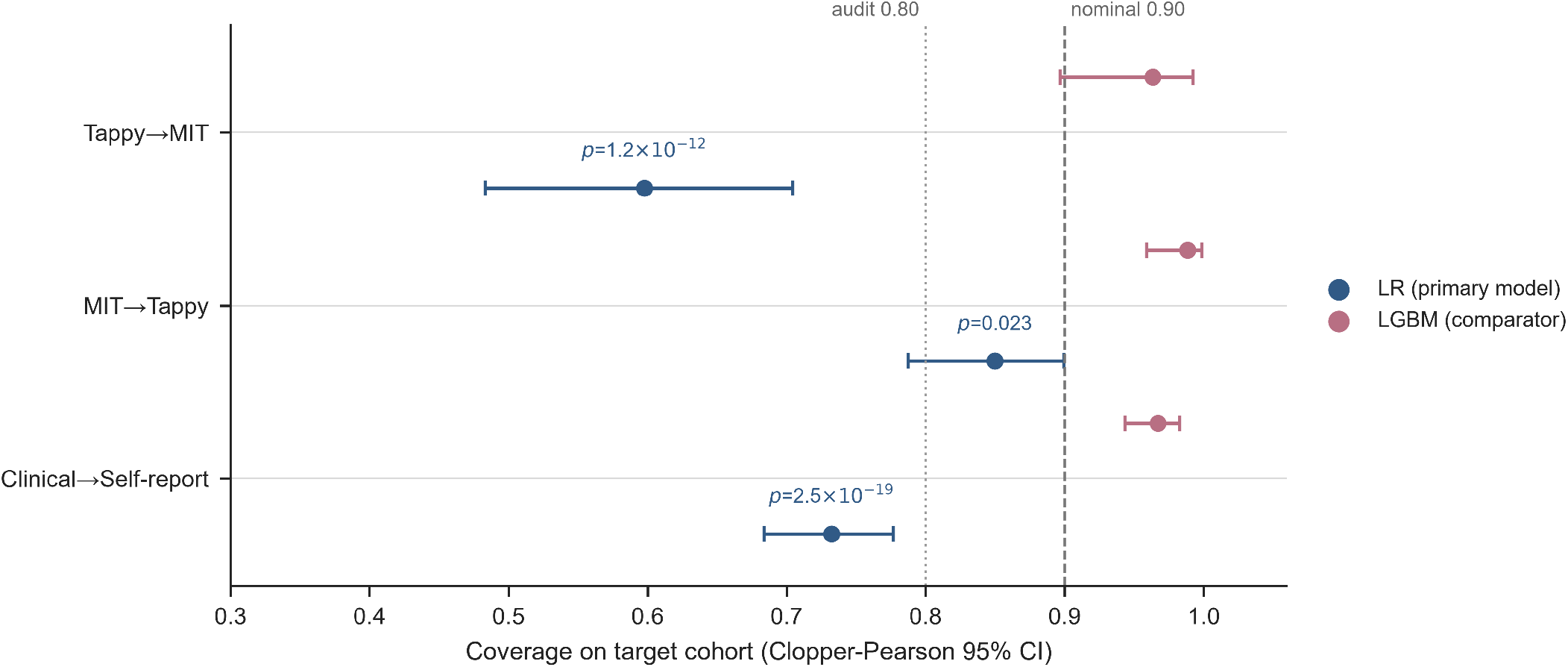
Marginal coverage and exact 95% Clopper—Pearson confidence intervals for the three primary transfer directions, comparing the primary model (LR) with the comparator model (LightGBM). Labels show Holm-adjusted p-values from one-sided exact binomial tests of H0: coverage ≥ 0.90 for the primary model; all three null hypotheses were rejected at family-wise error rate 0.10. Audit classifications follow the confidence-interval decision rule defined in the Methods and are tabulated with complete numerical results in Supplementary Table S5.

The mean set size across the three primary directions ranged from 1.11 to 1.53, the singleton rate ranged from 0.47 to 0.89, and the corresponding full-label-set output rates were 11%, 53%, and 30%, respectively. The complete audit matrix for all 12 ordered transfer directions is provided in Supplementary Table S2.

### 3.3. Coverage-discrimination decoupling

Across the 12 descriptive transfer directions, the AUC of the primary model ranged from 0.55 to 0.81 (mean across 50 seeds), and marginal coverage ranged from 0.58 to 0.91 (Supplementary Fig. S1). AUC deviation from the development-domain reference and coverage deviation from the nominal level did not vary in tandem across directions (Figure 2). Relative to the shared MIT development-domain benchmark, the primary-model AUC was 0.005-0.026 higher in three directions; two of these still had coverage more than 5 percentage points below 0.90. The other nine directions had AUC below the shared benchmark and coverage below the nominal level. In the reverse transfer pair, Tappy→MIT had higher mean AUC than MIT→Tappy (0.672 vs. 0.586) but a larger coverage deficit (23.1 vs. 4.7 percentage points; 50-seed means).

**Figure 2.**
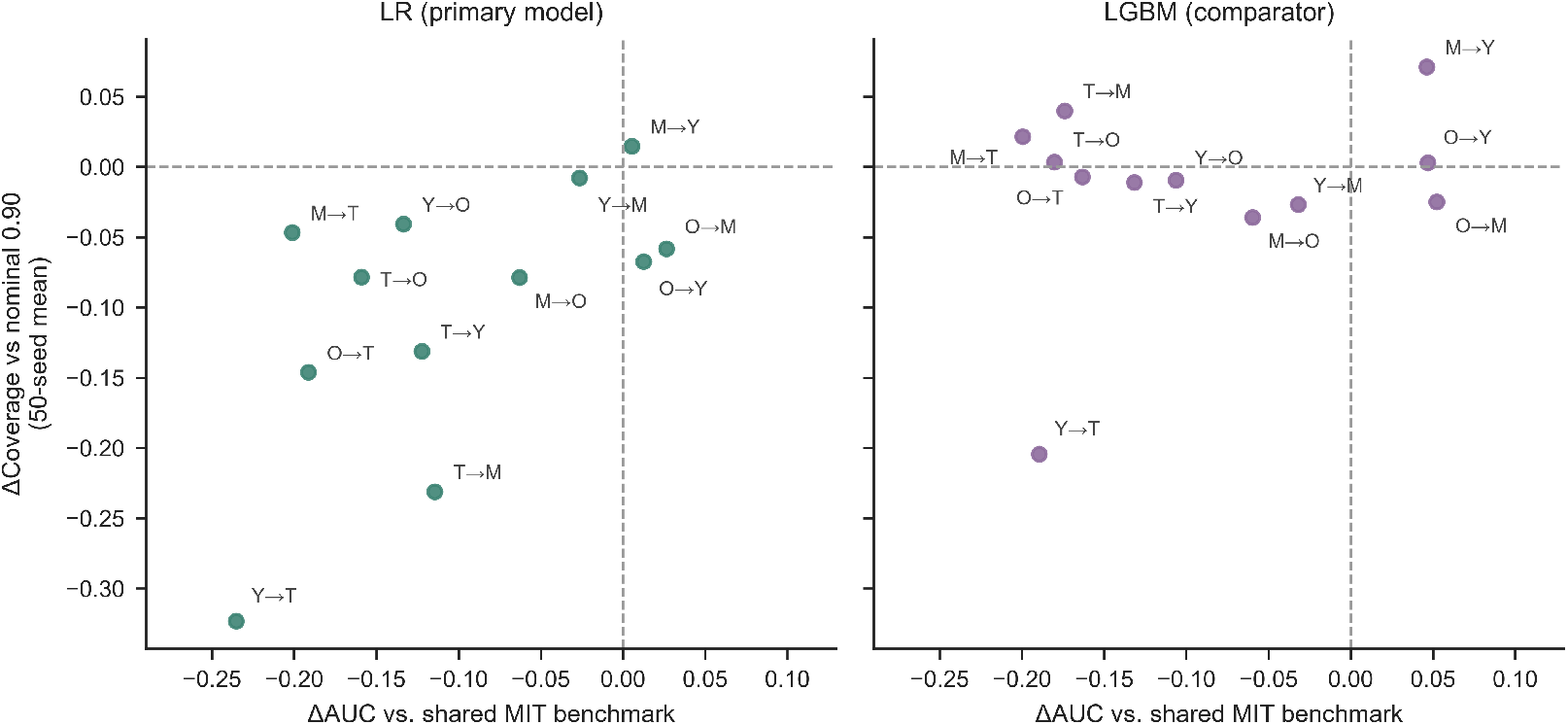
Deviation of AUC from the shared MIT development-domain benchmark versus deviation of coverage from the nominal level of 0.90, across the 12 descriptive transfer directions. Left panel: primary model (LR). Right panel: comparator (LGBM). Point labels abbreviate datasets (T, Tappy; M, MIT; Y, TyPD; O, Online English). Only 3 of the 12 directions exceeded the shared MIT benchmark (LR panel), and 2 of these had coverage more than 5 percentage points below nominal; deviations in the two dimensions did not co-vary.

The cross-model comparat or provided complementary evidence. Light-GBM had lower AUC than the primary model in all 12 directions (0.526− 0.778), while marginal coverage was higher in 11 of these directions. In the three primary directions, coverage reached 0.963-0.988, accompanied by larger prediction sets and lower singleton rates (mean set size 1.92-1.97; singleton rate 0.03-0.08).

### 3.4. Class-conditional coverage

In the Tappy →MIT direction, PD conditional coverage was 0.214 [0.103, 0.368] (n = 42), and HC conditional coverage was 1.000 (n = 40). Among the 42 participants with PD, 33 (78.6%) were not covered, and 34 (81.0%) received singleton prediction sets (Figure 3). Only 9 participants with PD were covered, and at least 25 of the 33 uncovered participants received singleton sets containing only the HC label. The PD proportions in the three target cohorts were 51% (42/82), 77% (133/173), and 59% (216/366), respectively. In the two target cohorts with the highest PD proportions, the undercovered class was HC (MIT→Tappy 0.700, n = 40; clinical pool-self-reported 0.627, n = 150). In Tappy→MIT, which had the lowest PD proportion and a more balanced distribution between the two classes, the undercovered class was PD. Class-conditional coverage for the comparator model was nearly uniform (0.925-1.000).

**Figure 3.**
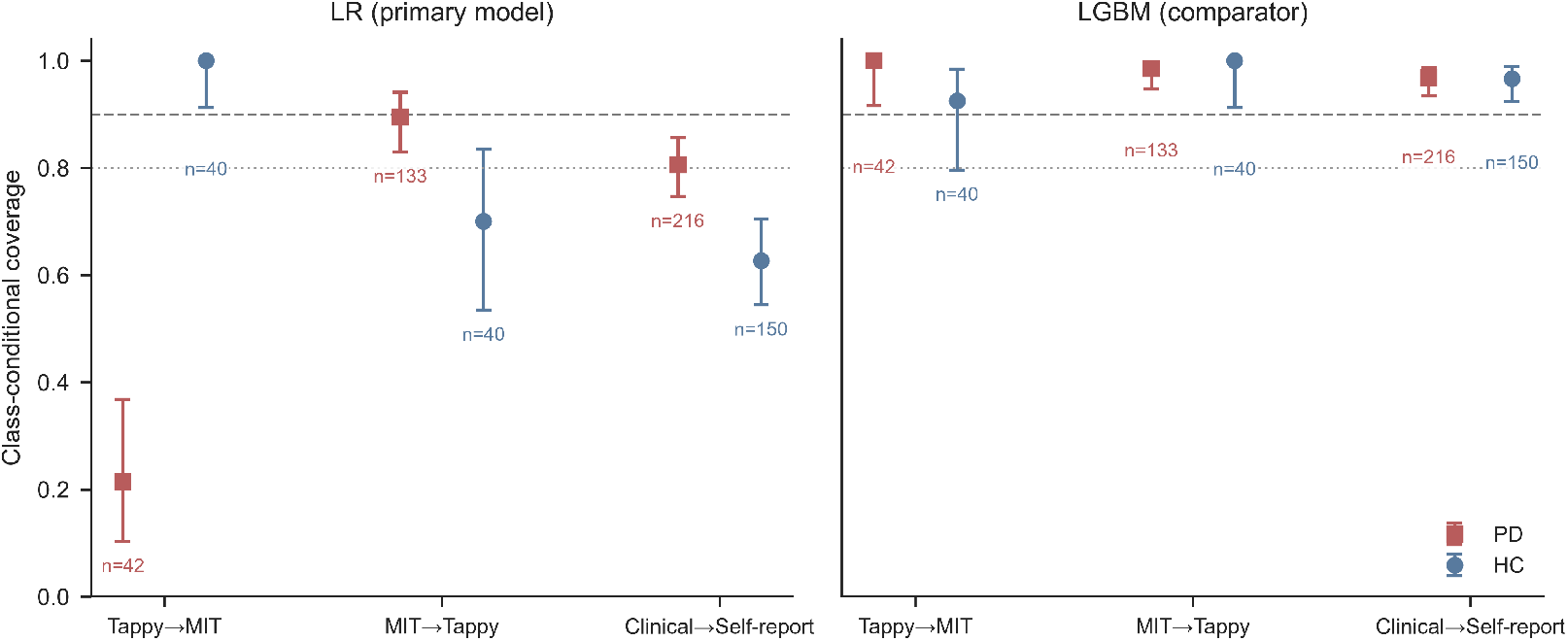
Class-conditional coverage with exact 95% confidence intervals for PD and HC in the three primary transfer directions. Left panel: primary model (LR). Right panel: comparator (LGBM). In Tappy→MIT, PD-conditional coverage was 0.214 while HC-conditional coverage was 1.000; the undercovered class differed across the three target cohorts.

### 3.5. Coverage recovery and efficiency costs

In the MIT→Tappy direction, coverage increased from 0.850 to 0.982 [0.947, 0.996] (9 target-domain labels, 5% budget, evaluated on the remaining 164 participants; Figure 4a). In the clinical pool→self-reported direction, coverage increased from 0.732 to 0.876 [0.837, 0.909] (18 target-domain labels, 5% budget). The lower bounds of the 95% confidence intervals for both directions reached the coverage audit threshold. The recovery effect of target-domain recalibration showed clear direction dependence, and Tappy→MIT followed a different pattern, with coverage increasing from 0.598 to 0.833 (16 target-domain labels, 20% budget) and reaching 0.877 [0.763, 0.949] at the 30% budget (25 target-domain labels), while the lower bound of the confidence interval remained below 0.80 and the residual PD conditional coverage deficit persisted (0.759, with HC at 1.000). Recalibration pooled across protocols improved marginal coverage but did not eliminate the class-conditional coverage gap. When fewer than 9 target-domain labels were used, k = [(n_cal_+1)(1-α)] exceeded the calibration sample size, and the prediction set degenerated to the full label set.

**Figure 4.**
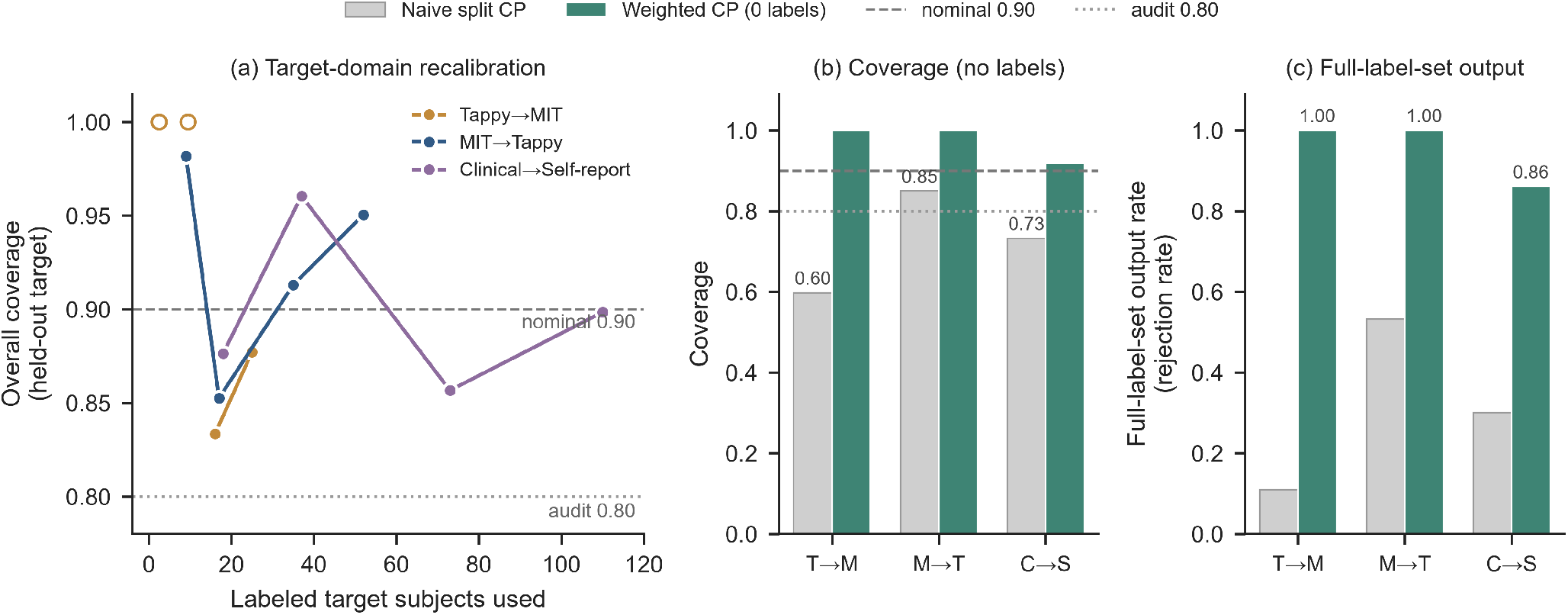
Coverage recovery and efficiency costs. (a) Overall coverage on held-out target subjects as a function of the number of labeled target subjects used for target-domain recalibration across the three primary directions; nine and 18 labels raised the confidence-interval lower bound above the 0.80 audit threshold in MIT→Tappy and the clinical pool→self-report direction, whereas Tappy→MIT remained below 0.80 at the 30% budget. (b) Coverage on held-out target subjects for naive split CP and weighted CP without target labels, across the three primary directions. (c) Full-label-set output rate (rejection rate) for the same comparison; weighted CP restored marginal coverage to 0.918–1.000 but returned the full label set for 86–100% of subjects. Panel (a) requires target-domain labels; panels (b) and (c) use unlabeled distributional information only.

Without increasing target-domain labeling, we further evaluated the feasibility of recovery using unlabeled distribution al information from the target domain. Weighted CP increased marginal coverage in the three primary directions to 0.918 1.000, but returned the full label set for 86 – 100% of participants (full-label-set output rate 0.86 – 1.00; Figure 4b, c), indicating that the recovery of marginal coverage was accompanied by a substantial loss in prediction-set effciency.

In addition to target-domain labels and unlabeled distributional information, the acquisition protocol provides another target-domain stratification factor that can be identified in advance. After calibration quantiles were calculated separately for each acquisition protocol, coverage in the target cohort pooled across the two protocols was 0.836 [0.793, 0.874] at a 5% labeling budget, and the lower bound of the confidence interval remained below the coverage audit threshold.

### 3.6. Stress tests of alternative shift mechanisms

Controlled stress tests were conducted in the least severely affected primary direction, MIT→Tappy. In the synthetic dose-response experiment, dose-dependent HC→PD label flipping caused HC conditional coverage to decrease monotonically from 0.700 to 0.425 while PD-side coverage remained at 1.000, whereas PD→HC and symmetric flipping did not collapse the opposite class (Supplementary Fig. S2): calibration-side label noise can produce one-sided, direction-dependent class-conditional collapse. However, none of the three manipulation scenarios reproduced the observed pattern of MIT→Tappy within the practical-equivalence tolerances; two scenarios reduced HC conditional coverage to 0.025 and 0.000, far below the observed 0.700. BBSE approximately recovered the target prevalence in constructed label-shift scenarios, whereas in Tappy→MIT it estimated a PD proportion of 0.000 against an observed 0.512 (the direction with the most severe audit failure), which did not support pure label shift as the dominant explanation for the observed coverage failure. Complete dose-response and scenario results are provided in Supplementary Fig. S2.

### 3.7. Robustness and subgroup analyses

Sensitivity analyses left the main findings unchanged in all three primary directions (Supplementary Table S6). When *α* was raised to 0.20, coverage ranged from 0.522 to 0.705; reclassifying the three participants who reported tremor only under either alternative label assignment shifted coverage by at most +0.073 (Tappy→MIT upper bound 0.771 when classified as HC, still below 0.90); re-aggregating sessions by quarter did not change the conclusions, and no monotonic gradient was observed across feature levels. For MIT→Tappy, the primary upper confidence bound (0.899) was close to the nominal coverage level of 0.90, and the mean coverage across 50 random seeds was 0.853 ± 0.058 (standard deviation; range 0.630–0.954). The power analysis is presented in Supplementary Table S3.

As a descriptive analysis, sex stratification was possible only within the Tappy target cohort (88 men and 85 women): coverage was lower in men than in women by 8.7 percentage points in MIT→Tappy (0.807 vs. 0.894) and by 12.8 percentage points in the clinical pool→self-reported direction (0.625 vs. 0.753).

## 4. Discussion

Discrimination and conformal coverage across four publicly available typing datasets and 12 ordered transfer directions showed asynchronous changes under distribution shift, and changes in one dimension could not be reliably inferred from the other. The comparator model further demonstrated the same separation. LightGBM had lower AUC than logistic regression in all 12 directions, but higher marginal coverage in 11 of them. This finding was consistent with the central hypothesis, indicating that coverage degradation and discrimination degradation constitute two separable dimensions of deployment reliability, and that external validation based on AUC alone is insufficient to characterize deployment reliability[25].

This decoupling reflects the different properties [17] measured by the two metrics. AUC measures pairwise ranking quality; conformal coverage depends on the calibration-score distribution and on exchangeability between calibration and target data. Distribution shift may simultaneously alter class separability, model confidence, the distribution of calibration scores, and class composition, but these changes do not necessarily have consistent effects on ranking performance and prediction set validity.

Marginal coverage provides an overall frequency guarantee but does not guarantee coverage separately for each class. Even when marginal coverage passes the audit, it is still necessary to determine which class is affected by the failure. Tappy→MIT is the clearest example. The class affected by undercoverage also differed across the three target cohorts, with undercoverage occurring in the HC class in the two target cohorts with PD proportions of 77% and 59%. In Tappy→MIT, where the two classes were nearly balanced, undercoverage occurred on the PD side. Thus, marginal coverage can detect a decline in overall validity but cannot identify its class-specific structure, making class-conditional coverage a necessary complementary diagnostic for deployment audits.

The stress tests indicate what the candidate mechanisms can and cannot explain. Label noise can produce one-sided, direction-dependent class-conditional collapse, yet the collapse side in the actual Tappy→MIT transfer (the PD side) differs from what calibration-side HC→PD flipping produces, and none of the three manipulation scenarios reproduced the observed pattern within the practical-equivalence tolerances. BBSE approximately recovered the target prevalence in constructed label-shift scenarios but not in Tappy→MIT; the estimates did not support pure label shift as the dominant explanation for the most severe failure.

Coverage recovery showed that different transfer directions required different calibration resources. Target-domain labeling was the most direct strategy: the label budget required to reach the audit threshold varied across directions, and Tappy→MIT did not reach it even at the 30% budget, with the PD conditional coverage deficit persisting. The effectiveness of target-domain recalibration is therefore direction-specific, and no fixed labeling budget applies across sites. Weighted CP [16] restored coverage without target-domain labels but returned the full label set (Figure 4b, c). Protocol-conditional calibration did not clear the audit threshold. Coverage recovery is therefore not simply a matter of success or failure, but involves trade-offs among coverage validity, access to target-domain information, and prediction-set efficiency.

These results indicate that cross-dataset deployment audits cannot be conducted using a single performance metric. AUC can reveal whether discrimination is maintained, marginal coverage can assess overall validity, class-conditional and subgroup coverage can identify failure patterns, and recovery experiments can quantify the target-domain information required to achieve the target coverage and the associated efficiency costs.

This study has several limitations. The stress tests examined whether candidate mechanisms could reproduce the observed patterns under controlled conditions, but they cannot establish that these mechanisms caused the patterns in the data; comparisons across transfer directions remain observational. The primary analyses relied on a single calibration split, with the results across 50 random seeds reported only descriptively. For MIT→Tappy, the upper confidence bound was 0.899, close to the nominal coverage level of 0.90, so the audit classification for this direction may be sensitive to the calibration split. Δ AUC was computed against the MIT development-domain benchmark for each model; for directions from other source datasets, deviations partly reflect source-domain model differences rather than transfer effects alone. Target-domain recalibration samples were drawn with class-stratified sampling; in prospective deployment, labels would be obtained for randomly selected participants rather than class-balanced samples, so the reported label budgets characterize this design rather than a deployable minimum. Sex-stratified analyses were possible only for transfer directions whose target cohorts included Tappy participants, because MIT and OE do not provide demographic data. The four datasets also differed in diagnostic label source (clinical diagnosis or self-report) and data-collection platform, both of which are part of the deployment heterogeneity examined here. The extent to which the findings apply to other typing modalities or populations remains to be evaluated. The study used research-grade classifiers on public datasets and did not assess their validity or utility for clinical screening.

Future studies should extend the audit to other typing modalities and collection settings and prospectively assess whether coverage-aware deployment reporting informs model selection and monitoring decisions.

## 5. Conclusion

The audit across four publicly available typing datasets showed that AUC and conformal coverage changed asynchronously during cross-dataset transfer. Class-conditional coverage further revealed class-asymmetric failures that were not fully captured by marginal coverage, and the affected class varied across target cohorts. The effectiveness of coverage recovery varied by transfer direction and recovery strategy, involving trade-offs between target-domain information requirements and prediction-set efficiency. Therefore, deployment evaluation should report marginal and class-conditional coverage by target cohort in addition to AUC and take into account the resources required for coverage recovery and associated prediction-set efficiency.

## Data Availability

All datasets are publicly available: MIT-CSXPD (PhysioNet, https://physionet.org/content/nqmitcsxpd/1.0.0/), Tappy (PhysioNet, https://physionet.org/content/tappy/1.0.0/), TyPD (i-PROGNOSIS touch screen typing data; Zenodo https://zenodo.org/records/2571623), and Online English (OSF repository https://osf.io/ew34b/). The aggregate analysis outputs generated in this study are archived at https://doi.org/10.5281/zenodo.23553162

## CRediT authorship contribution statement

Haotian Guang: Conceptualization, Methodology, Software, Formal analysis, Data curation, Writing – original draft.

Zijuan Zhang: Validation, Writing – review & editing.

Jiale Qu: Validation, Writing – review & editing.

Jiaqiang Wu: Supervision, Writing – review & editing.

Jiayuan Ma: Formal analysis, Writing – review & editing.

## Declaration of competing interest

The authors declare that they have no known competing financial interests or personal relationships that could have appeared to influence the work reported in this paper.

### Ethics approval and consent to participate

This was a retrospective secondary analysis of publicly available, de-identified datasets; no new human data were collected. No ethics approval was required, and consent was not applicable (consent for the original datasets is documented in their source publications).

### Clinical trial number

Not applicable.

## Data availability

All datasets are publicly available: MIT-CSXPD (PhysioNet, nqmitcsxpd v1.0.0), Tappy (PhysioNet, tappy v1.0.0), TyPD (i-PROGNOSIS touchscreen typing data; Zenodo record 2571623), and Online English (OSF repository ew34b). The aggregate analysis outputs generated in this study are archived at https://doi.org/10.5281/zenodo.23553162.

## Code availability

The underlying code supporting the findings of this study is available at https://github.com/haotian-bio/typing-pd-conformal-audit.

### Declaration of generative AI and AI-assisted technologies in the manuscript preparation process

During the preparation of this work the authors used GPT-6 Astra (Ope-nAI) in order to assist with analysis code implementation and to revise and refine manuscript text drafted by the authors. After using this tool, the authors reviewed and edited the content as needed and take full responsibility for the content of the published article. The authors verified all numerical results against the archived analysis outputs.

## Acknowledgements

The authors thank the producers of the publicly available datasets used in this study: the neuroQWERTY/MIT-CSXPD team (PhysioNet), the i-PROGNOSIS consortium (TyPD), the Tappy dataset contributors, and the Online English dataset authors.

