## Supplementary Figure1, 2 for "A Cross-Dataset Audit of Discrimination and Conformal Prediction Coverage in Keystroke-Based Parkinson’s Disease Classification"

**Supplementary Fig. S1. Mean test AUC of the primary model across the 12 ordered cross-dataset transfer directions, averaged over 50 random seeds. Rows represent source datasets and columns represent target datasets. Mean AUC ranged from 0.552 to 0.813 and was below the in-domain reference in 9 of the 12 directions. Diagonal cells represent same-dataset pairs and are not included in the cross-dataset audit.**

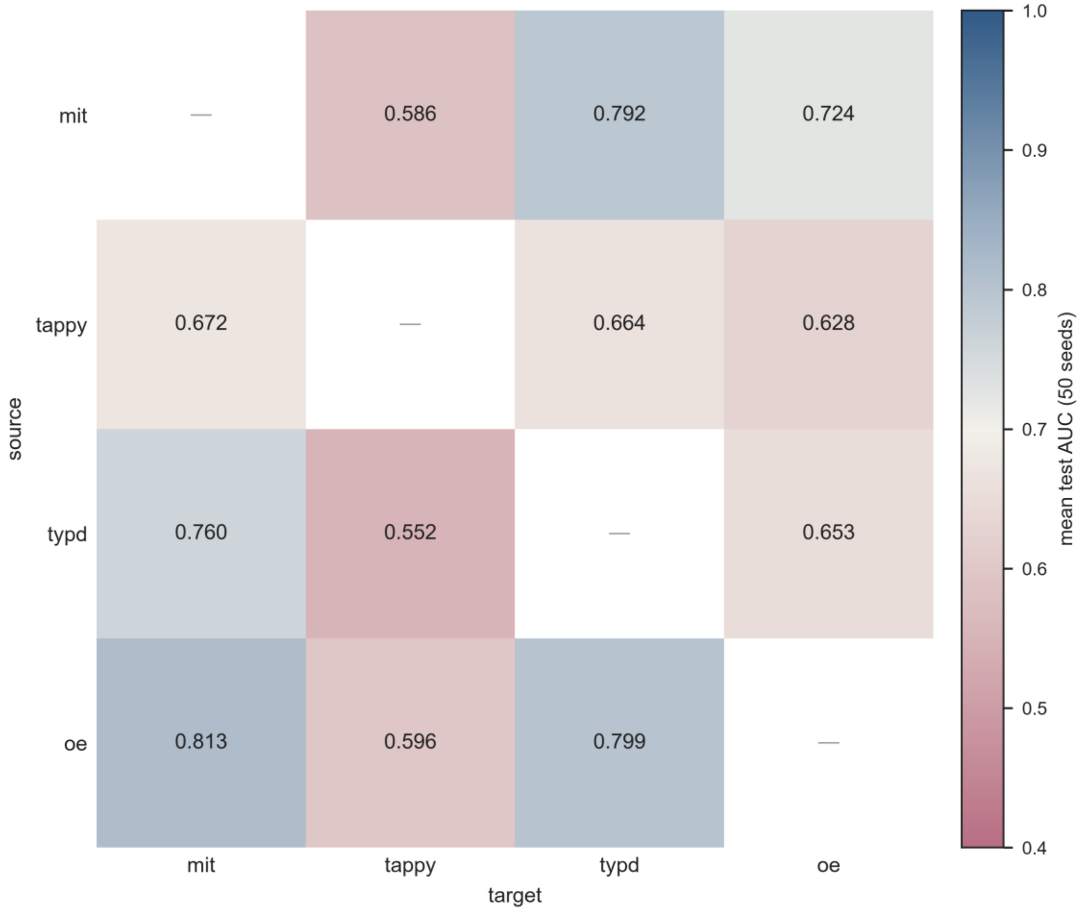


**Supplementary Fig. S2. Controlled stress tests in the MIT→Tappy direction. (a) Calibration-only label flipping (theory-aligned scope). (b) The same under the train+calibration scope. (c) Three-arm test on the real transfer: PD- and HC-conditional coverage for the observed pattern, prevalence re-matching, prevalence-preserving flips, and their combination. Label flipping produced one-sided class-conditional collapse, but none of the three manipulation scenarios reproduced the observed pattern within the practical-equivalence tolerances.**

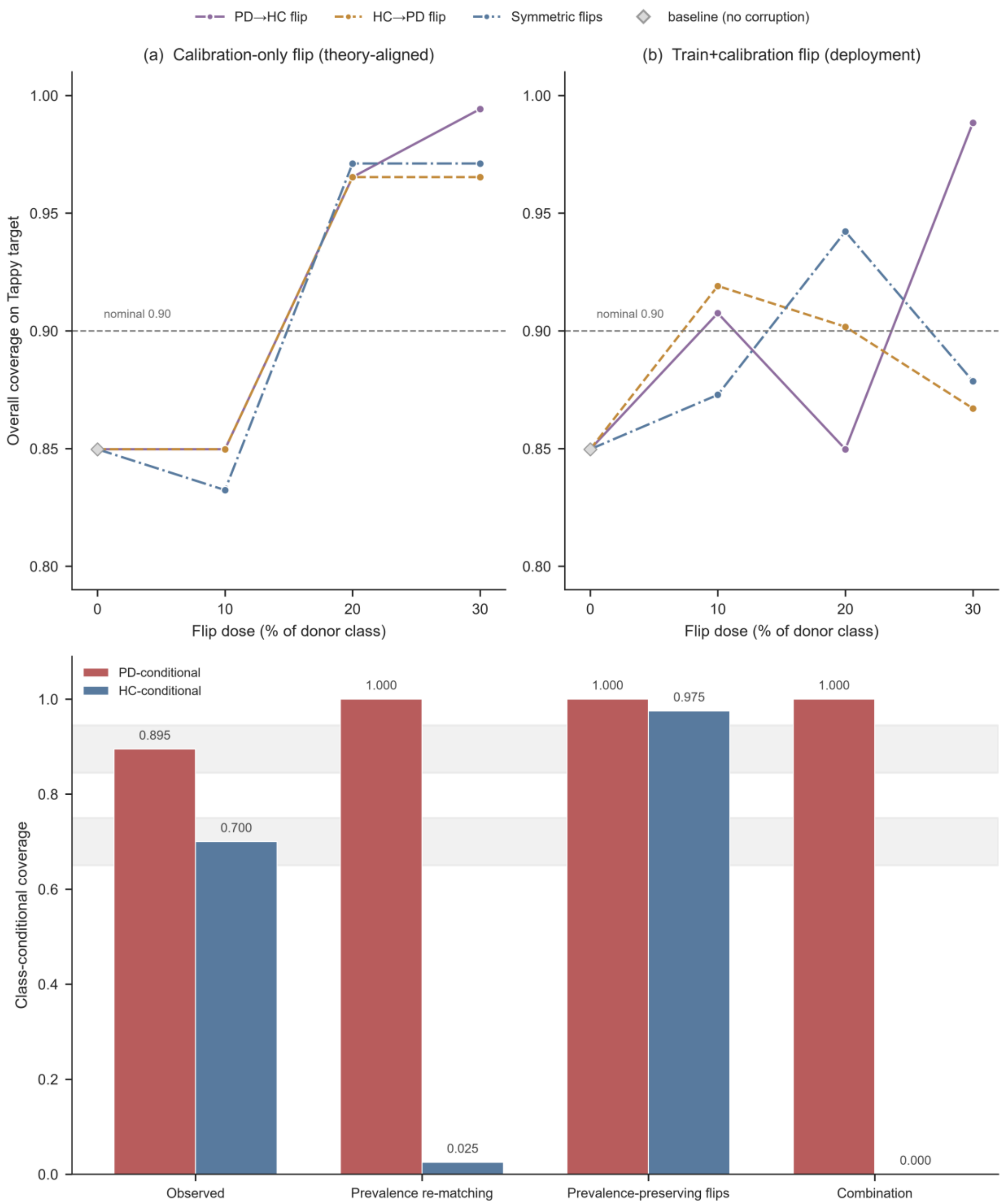


(c) Three-arm real-transfer test (shaded = ±5 pp practical-equivalence tolerance; not a CI)
