## Supplementary Table1 for "A Cross-Dataset Audit of Discrimination and Conformal Prediction Coverage in Keystroke-Based Parkinson’s Disease Classification"

### Supplementary Table S1. Data cleaning rules and sample attrition

All analysis metrics use the digraph, an ordered pair of consecutive keystrokes, as the basic unit of computation. Three timing quantities are derived per digraph: hold is the key press duration, latency is the time from the release of the previous keystroke to the press of the current keystroke, and flight is the interval between two consecutive keystrokes. A session is one continuous typing run: visit files for MIT, monthly files for Tappy, visits for TyPD, and single continuous entries for OE. Data cleaning did not reference participants' diagnostic labels at any step and relied solely on the quality of the typing data.

#### Panel A. Cleaning rules (applied in this order)

| Step | Rule | Applies to |
|---|---|---|
| S0 | Parsed press–release events on record. | All datasets |
| S1 | Digraph pool: parse-validated events with complete press and release timestamps; the first digraph of each session has no predecessor (no_predecessor: MIT 116, Tappy 0, TyPD 338, OE 3,442). | All |
| S2 | Universal timing: remove digraphs with hold or latency ≤ 0 s or > 10 s. | All |
| S3 | Fixed-text rule: remove digraphs with flight > 3 s. | MIT, TyPD, OE (fixed-text transcription); Tappy is free text and exempt |
| S4 | Visit selection: retain each participant's first clinical visit only (31 MIT sessions removed). | MIT |
| S5 | Session rules: typing rate < 20 characters per minute; sessions with < 100 valid digraphs removed. | Rate rule: MIT, TyPD, OE; valid-digraph rule: all |
| S6 | Person rule: participants with < 300 valid digraphs excluded. | All |
| S7 | Label rules: participants without diagnosis labels excluded; labeled participants whose records contain no parseable events never enter the pipeline; three Tappy participants reporting tremor without a PD diagnosis excluded from the primary analysis (re-inclusion under each label assignment reported as a sensitivity analysis). | All |
| S8 | Final analysis cohort: 475 participants (PD 270 / HC 205), corresponding to Table 1. | All |

#### Panel B. Sample attrition by step (participants / sessions / digraphs remaining)

| Step | MIT | Tappy | TyPD | Online English |
|---|---|---|---|---|
| S0 | 85 / 116 / 171,973 | 266 / 621 / 9,316,858 | 33 / 33 / 26,343 | 232 / 3,451 / 437,915 |
| S1 | 85 / 116 / 171,857 | 266 / 621 / 9,316,025 | 33 / 33 / 26,005 | 232 / 3,451 / 434,464 |
| S2 | 85 / 116 / 159,030 | 266 / 621 / 9,301,264 | 33 / 33 / 25,893 | 232 / 3,451 / 351,761 |

| | | | | |
|---|---|---|---|---|
| S3 | 85 / 116 / 155,408 | n/a | 33 / 33 / 24,123 | 232 / 3,451 / 350,272 |
| S4 | 85 / 116 / 155,408 | n/a | n/a | 232 / 3,406 / 350,272 |
| S5 | 85 / 82 / 112,014 | 266 / 560 / 9,298,974 | 33 / 27 / 20,822 | 232 / 1,891 / 237,565 |
| S6 | 82 / 82 / 112,014 | 205 / 538 / 9,295,008 | 27 / 27 / 20,822 | 193 / 1,861 / 234,368 |
| S7–S8 | 82 (42/40) | 173 (133/40) | 27 (12/15) | 193 (83/110) |

Notes. Counts are participants / sessions / digraphs remaining after each step; S7–S8 shows participants (PD/HC).

(1) Session counts up to S3 include sessions with no surviving digraphs; from S4, sessions are counted among the pooled digraphs (45 Online English sessions had no surviving digraphs).

(2) At S7, 29 participants without labels and 3 tremor-only participants were removed from the 205 Tappy survivors; a further 10 labeled Tappy participants never entered the pipeline because their records contained no parseable events (they are not included in the counts above).

(3) For TyPD, sessions are clinical visits (338 trials in total).
