## Supplementary Table2 for "A Cross-Dataset Audit of Discrimination and Conformal Prediction Coverage in Keystroke-Based Parkinson’s Disease Classification"

**Supplementary Table S2. Complete audit matrix for the 12 ordered transfer directions (discrimination, coverage, and prediction-set efficiency; the primary model (LR) and the comparator model (LightGBM); 50 random seeds; AUC and coverage as mean ± SD)**

| Direction | Level | N src | N tgt | LR AUC | LR coverage | LR PD cond. | LR HC cond. | LR set size | LR singleton | LR full set | LGBM AUC | LGBM coverage | LGBM PD cond. | LGBM HC cond. | LGBM set size | LGBM singleton | LGBM full set |
|---|---|---|---|---|---|---|---|---|---|---|---|---|---|---|---|---|---|
| MIT→Tappy | 2 | 82 | 173 | 0.586 ± 0.01 | 0.853 ± 0.06 | 0.942 | 0.557 | 1.42 | 0.585 | 0.415 | 0.526 ± 0.03 | 0.922 ± 0.05 | 0.958 | 0.800 | 1.75 | 0.254 | 0.746 |
| MIT→TyPD | 1 | 82 | 27 | 0.792 ± 0.02 | 0.915 ± 0.06 | 0.895 | 0.931 | 1.46 | 0.538 | 0.462 | 0.772 ± 0.06 | 0.971 ± 0.05 | 0.937 | 0.999 | 1.77 | 0.225 | 0.775 |
| MIT→OE | 3 | 82 | 193 | 0.724 ± 0.02 | 0.821 ± 0.08 | 0.984 | 0.699 | 1.51 | 0.490 | 0.510 | 0.666 ± 0.02 | 0.864 ± 0.07 | 0.902 | 0.835 | 1.58 | 0.419 | 0.581 |
| Tappy→MIT | 2 | 173 | 82 | 0.672 ± 0.04 | 0.669 ± 0.14 | 0.356 | 0.997 | 1.23 | 0.771 | 0.229 | 0.552 ± 0.09 | 0.940 ± 0.08 | 0.999 | 0.879 | 1.87 | 0.127 | 0.873 |
| Tappy→TyPD | 1 | 173 | 27 | 0.664 ± 0.11 | 0.769 ± 0.11 | 0.555 | 0.940 | 1.34 | 0.658 | 0.342 | 0.594 ± 0.15 | 0.889 ± 0.11 | 1.000 | 0.800 | 1.79 | 0.212 | 0.788 |
| Tappy→OE | 2 | 173 | 193 | 0.628 ± 0.03 | 0.822 ± 0.10 | 0.676 | 0.932 | 1.51 | 0.489 | 0.511 | 0.545 ± 0.04 | 0.903 ± 0.10 | 0.999 | 0.831 | 1.82 | 0.178 | 0.822 |
| TyPD→MIT | 1 | 27 | 82 | 0.760 ± 0.04 | 0.892 ± 0.08 | 0.800 | 0.988 | 1.56 | 0.435 | 0.565 | 0.694 ± 0.06 | 0.873 ± 0.10 | 0.775 | 0.977 | 1.59 | 0.410 | 0.590 |
| TyPD→Tappy | 1 | 27 | 173 | 0.552 ± 0.01 | 0.577 ± 0.19 | 0.453 | 0.989 | 1.42 | 0.579 | 0.421 | 0.536 ± 0.03 | 0.696 ± 0.22 | 0.605 | 0.997 | 1.59 | 0.411 | 0.589 |
| TyPD→OE | 1 | 27 | 193 | 0.653 ± 0.04 | 0.859 ± 0.10 | 0.704 | 0.976 | 1.56 | 0.432 | 0.568 | 0.619 ± 0.04 | 0.891 ± 0.13 | 0.764 | 0.986 | 1.68 | 0.319 | 0.681 |
| OE→MIT | 3 | 193 | 82 | 0.813 ± 0.02 | 0.842 ± 0.05 | 0.701 | 0.989 | 1.34 | 0.658 | 0.342 | 0.778 ± 0.03 | 0.875 ± 0.04 | 0.757 | 0.999 | 1.55 | 0.453 | 0.547 |
| OE→Tappy | 2 | 193 | 173 | 0.596 ± 0.01 | 0.754 ± 0.13 | 0.755 | 0.748 | 1.41 | 0.588 | 0.412 | 0.562 ± 0.02 | 0.893 ± 0.08 | 0.914 | 0.821 | 1.66 | 0.337 | 0.663 |
| OE→TyPD | 1 | 193 | 27 | 0.799 ± 0.01 | 0.833 ± 0.06 | 0.922 | 0.761 | 1.36 | 0.639 | 0.361 | 0.773 ± 0.06 | 0.903 ± 0.07 | 0.782 | 1.000 | 1.59 | 0.407 | 0.593 |

(1) AUC is the participant-level test AUC on the target dataset; coverage, PD/HC conditional coverage, mean set size, singleton rate, and full-set output rate are means over the 50 random seeds (SD reported for AUC and coverage). Conditional coverage is coverage among PD and among HC participants, respectively.

(2) Level is the effective feature level of the direction (the smaller of the source and target ceilings); N src / N tgt are the cohort sizes. The full-set output rate equals the doubleton rate (in binary classification, a prediction set containing both classes) and is the abstention fraction; the empty-set rate is 0.

(3) The 12 ordered directions are descriptive; statistical tests for the three primary directions (including the pooled clinical-pool→self-report direction) are reported in manuscript Table 2. Confidence intervals are defined only on the primary single split and are not reported here.

(4) Discrimination was not tested inferentially; the 50-seed AUC summaries characterize stability across splits. The direction-level joint interpretation of AUC and coverage is presented in the manuscript and Figure 3.
