## Supplementary Table3 for "A Cross-Dataset Audit of Discrimination and Conformal Prediction Coverage in Keystroke-Based Parkinson’s Disease Classification"

**Supplementary Table S3. Power analysis: distribution of 0.80 audit-rule verdicts as a function of true coverage (50,000 simulations per cell)**

Panel A. Planned sample-size scenarios

| Direction | Target-cohort n | True coverage | P(Compatible) | P(Incompatible) | P(Undetermined) |
|---|---|---|---|---|---|
| Tappy→MIT | 85 | 0.90 | 0.658 | 0.020 | 0.321 |
| Tappy→MIT | 85 | 0.85 | 0.162 | 0.289 | 0.549 |
| Tappy→MIT | 85 | 0.80 | 0.016 | 0.745 | 0.239 |
| Tappy→MIT | 85 | 0.70 | 0.000 | 0.997 | 0.003 |
| MIT→Tappy | 227 | 0.90 | 0.981 | 0.019 | 0.000 |
| MIT→Tappy | 227 | 0.85 | 0.397 | 0.603 | 0.000 |
| MIT→Tappy | 227 | 0.80 | 0.013 | 0.987 | 0.000 |
| MIT→Tappy | 227 | 0.70 | 0.000 | 1.000 | 0.000 |
| Clinical pool→self-report | 457 | 0.90 | 0.983 | 0.017 | 0.000 |
| Clinical pool→self-report | 457 | 0.85 | 0.116 | 0.884 | 0.000 |
| Clinical pool→self-report | 457 | 0.80 | 0.000 | 1.000 | 0.000 |
| Clinical pool→self-report | 457 | 0.70 | 0.000 | 1.000 | 0.000 |
| MIT→TyPD | 33 | 0.90 | 0.144 | 0.013 | 0.842 |
| MIT→TyPD | 33 | 0.85 | 0.033 | 0.110 | 0.857 |
| MIT→TyPD | 33 | 0.80 | 0.006 | 0.335 | 0.659 |
| MIT→TyPD | 33 | 0.70 | 0.000 | 0.820 | 0.180 |

Panel B. Realized sample sizes (as in manuscript Table 1)

| Direction | Target-cohort n | True coverage | P(Compatible) | P(Incompatible) | P(Undetermined) |
|---|---|---|---|---|---|
| Tappy→MIT | 82 | 0.90 | 0.698 | 0.015 | 0.286 |
| Tappy→MIT | 82 | 0.85 | 0.195 | 0.242 | 0.562 |
| Tappy→MIT | 82 | 0.80 | 0.022 | 0.690 | 0.287 |
| Tappy→MIT | 82 | 0.70 | 0.000 | 0.995 | 0.005 |
| MIT→Tappy | 173 | 0.90 | 0.960 | 0.024 | 0.016 |
| MIT→Tappy | 173 | 0.85 | 0.388 | 0.528 | 0.085 |
| MIT→Tappy | 173 | 0.80 | 0.024 | 0.962 | 0.014 |
| MIT→Tappy | 173 | 0.70 | 0.000 | 1.000 | 0.000 |
| Clinical pool→self-report | 366 | 0.90 | 0.978 | 0.022 | 0.000 |
| Clinical pool→self-report | 366 | 0.85 | 0.175 | 0.825 | 0.000 |
| Clinical pool→self-report | 366 | 0.80 | 0.000 | 1.000 | 0.000 |
| Clinical pool→self-report | 366 | 0.70 | 0.000 | 1.000 | 0.000 |
| MIT→TyPD | 27 | 0.90 | 0.232 | 0.014 | 0.753 |
| MIT→TyPD | 27 | 0.85 | 0.072 | 0.099 | 0.829 |
| MIT→TyPD | 27 | 0.80 | 0.020 | 0.286 | 0.695 |
| MIT→TyPD | 27 | 0.70 | 0.001 | 0.743 | 0.256 |

Notes. (1) Each cell gives, over 50,000 simulations, the probability of each verdict under the 0.80 audit rule (Compatible: 95% CI lower bound ≥ 0.80; Incompatible: upper bound < 0.90; otherwise Undetermined).

(2) The first three directions are the primary directions; MIT→TyPD is a non-primary direction, included for reference.

(3) Panel B uses the realized cohorts: at true coverage 0.70, all three primary directions are classified as Incompatible with probability close to 1.00; at true coverage 0.90, MIT→Tappy and clinical-pool→self-report are classified as Compatible with probability ≥ 0.96.
