## Supplementary Table4 for "A Cross-Dataset Audit of Discrimination and Conformal Prediction Coverage in Keystroke-Based Parkinson’s Disease Classification"

**Supplementary Table S4. Datasets and analysis cohorts (participant counts; N (PD/HC)). Typing task and session rules are defined in the Methods of the main text; level is the effective feature level.**

| Dataset | Labels | Level | N (PD/HC) | Role |
|---|---|---|---|---|
| MIT-CSXPD | clinical | L3 | 82 (42/40) | Tappy→MIT target; MIT→Tappy source; clinical pool |
| Tappy Keystroke | self-report | L2 | 173 (133/40) | MIT→Tappy target; Tappy→MIT source; self-report pool |
| TyPD (i-PROGNOSIS) | clinical | L1 | 27 (12/15) | clinical pool; MIT→TyPD target |
| Online English | self-report | L3 | 193 (83/110) | self-report pool |
| Total | | | 475 (270/205) | |
