## Supplementary Table5 for "A Cross-Dataset Audit of Discrimination and Conformal Prediction Coverage in Keystroke-Based Parkinson’s Disease Classification"

**Supplementary Table S5. Primary coverage audit (α = 0.10, split CP, single primary split). Tests of H0: true coverage ≥ 0.90 under Holm correction (family-wise error control). LR = logistic regression (primary model); LGBM = LightGBM (comparator).**

| Direction | Model | K/n | Coverage (95% CI) | Adjusted p | Verdict |
|---|---|---|---|---|---|
| Tappy→MIT | LR | 49/82 | 0.598 (0.483–0.704) | $1.2 \times 10^{-12}$ | Incompatible |
| Tappy→MIT | LGBM | 79/82 | 0.963 (0.897–0.992) | — | — |
| MIT→Tappy | LR | 147/173 | 0.850 (0.788–0.899) | 0.023 | Incompatible |
| MIT→Tappy | LGBM | 171/173 | 0.988 (0.959–0.999) | — | — |
| Clinical pool→self-report | LR | 268/366 | 0.732 (0.684–0.777) | $2.5 \times 10^{-19}$ | Incompatible |
| Clinical pool→self-report | LGBM | 354/366 | 0.967 (0.943–0.983) | — | — |
