## Supplementary Table6 for "A Cross-Dataset Audit of Discrimination and Conformal Prediction Coverage in Keystroke-Based Parkinson’s Disease Classification"

**Supplementary Table S6. Sensitivity analyses (three primary transfer directions). Except for the anchor rows, each group of rows modifies a single setting; the anchor rows reproduce the primary-analysis values of Supplementary Table S5 for each direction.**

| Variant | Direction | K/n | Coverage (95% CI) |
|---|---|---|---|
| Anchor (primary analysis) | Tappy→MIT | 49/82 | 0.598 (0.483–0.704) |
| Anchor (primary analysis) | MIT→Tappy | 147/173 | 0.850 (0.788–0.899) |
| Anchor (primary analysis) | Clinical pool→self-report | 268/366 | 0.732 (0.684–0.777) |
| α raised to 0.20 | Tappy→MIT | 47/82 | 0.573 (0.459–0.682) |
| α raised to 0.20 | MIT→Tappy | 122/173 | 0.705 (0.631–0.772) |
| α raised to 0.20 | Clinical pool→self-report | 191/366 | 0.522 (0.469–0.574) |
| Tremor-only as PD | Tappy→MIT | 49/82 | 0.598 (0.483–0.704) |
| Tremor-only as PD | MIT→Tappy | 147/176 | 0.835 (0.772–0.887) |
| Tremor-only as PD | Clinical pool→self-report | 268/369 | 0.726 (0.678–0.771) |
| Tremor-only as HC | Tappy→MIT | 55/82 | 0.671 (0.558–0.771) |
| Tremor-only as HC | MIT→Tappy | 150/176 | 0.852 (0.791–0.901) |
| Tremor-only as HC | Clinical pool→self-report | 271/369 | 0.734 (0.686–0.779) |
| Sessions by quarter | Tappy→MIT | 49/82 | 0.598 (0.483–0.704) |
| Sessions by quarter | MIT→Tappy | 147/173 | 0.850 (0.788–0.899) |
| Sessions by quarter | Clinical pool→self-report | 268/366 | 0.732 (0.684–0.777) |
| 50 seeds (mean, range) | Tappy→MIT | — | 0.669 (0.488–0.976) |
| 50 seeds (mean, range) | MIT→Tappy | — | 0.853 (0.630–0.954) |
| 50 seeds (mean, range) | Clinical pool→self-report | — | 0.852 (0.691–0.937) |
